# Impact of the additional/booster dose of COVID-19 vaccine against severe disease during the epidemic phase characterized by the predominance of the Omicron variant in Italy, December 2021 - May 2022

**DOI:** 10.1101/2022.04.21.22273567

**Authors:** Francesco Branda

## Abstract

Vaccine effectiveness (VE) against emerging SARS-CoV-2 variants is a growing issue. The aim of this work is to evaluate the impact of the COVID-19 additional/booster dose against COVID-19-related symptoms, hospitalization, and death using surveillance data (December 2021-May 2022) from the Italian National Institute of Health (ISS). As of 18 May 2022, for people fully vaccinated with three doses, VE was 87.8% (95% confidence interval (CI): 87.6-88.0), whereas VE decreased from 71.5% (95% CI: 71.0–72.0) in those vaccinated > 120 days to 69.6% (95% CI: 68.3-70.8) within 91-120 days. Results support the importance of receiving a third dose of mRNA COVID-19 vaccine.

## Introduction

The development of effective and safe vaccines against severe acute respiratory syndrome coronavirus 2 (SARS-CoV-2) has been extraordinarily rapid, with less than a year elapsing between the sequencing of the new viral genome and the initiation of major vaccination campaigns in different parts of the world. However, several notable variants of SARS-CoV-2 (also known as *variants of concern* (VOCs)) continue to emerge, such as the Omicron variant, causing high rates of infection/re, a major impact on health care services, and a slowdown to the socio-economic system [1]. Several studies contributed to show the effectiveness of vaccination against symptomatic disease caused by the VOCs [2,3]. Other studies investigated the potential decline in vaccine-induced protective immunity [4,5,6], showing that protection against symptomatic disease wanes over time against symptomatic disease [7,8].

Regardless of the vaccine received as a 2nd dose, the mRNA vaccines provide a strong booster effect with low reactogenicity [9], so the competent authorities recommended either a BNT162b2 (Comirnaty) or a half dose (50 µg) of mRNA-1273 (Spikevax) vaccine to be given as a booster dose. In Italy, COVID-19 booster vaccines were introduced on 13 September 2021. The doses were initially offered only to those with suppressed immune systems, including AIDs patients and those on dialysis for renal failure, as well as cancer patients and transplant recipients. However, to contain rising cases, the eligibility criteria were quickly expanded to include care home workers, the over 80s, and health professionals by the end of September; over-60s by mid-October; over-40s by late November; and over-18s by the end of December. From February 1, 2022, the minimum period to receive the booster dose of vaccine is reduced from 5 to 4 months after the last dose.

The aim of this work is to evaluate the impact of COVID-19 additional/booster vaccines against COVID-19-related symptoms, hospitalization, and death in Italy, between December 28, 2021, and May 18, 2022, when Omicron was the dominant variant in circulation.

## Materials and Methods

The data used in this work are extracted from the bulletins of the Istituto Superiore di Sanità (ISS) starting from 28 December 2021^1^ and then made available in machine-readable format on the GitHub platform^2^.

The estimates of vaccine effectiveness (VE) reported in the weekly bulletin of ISS^1^ are analyzed to evaluate the impact the additional/booster doses against severe illness during the epidemic phase characterized by the predominance of the Omicron variant. Such estimates are calculated using generalized linear random-effects model with Poisson distribution, considering the number of events per day as the dependent variable, vaccination status as the independent variable, 10-year age groups and weekly regional incidence as adjustment variables, and including region of administration as a random effect.

Using this model, it is possible to estimate the relative risk (RR), i.e., the ratio of the risks for an event for the exposure group to the risks for the non-exposure group. Moreover, the number needed to treat (NNT), i.e., the number of people who need to be treated to prevent one additional adverse outcome from a disease, is used to evaluate the impact of the VE. For more details, see Supplementary material.

### Results of the epidemiological investigation

As of 18/05/2022, 137,446,955 doses have been administered (48,813,521 first doses, 47,923,845 second/single doses and 39,496,795 third doses) [11]. Table 2 summarizes the results of the epidemiological investigation. Overall, 7,330,719 cases were reported among the unvaccinated, 5,269,976 cases among those vaccinated with 2 doses within 120 days, 10,691,343 cases among those vaccinated with 2 doses for more than 120 days, and 15,512,602 cases among those vaccinated with an additional/booster dose (see Table 1). Since it is not possible to compare the absolute numbers of events in the different vaccination status within the same age group because they refer to different populations, it was necessary to calculate the incidence rate (IR), i.e., the number of new cases of a disease divided by the number of persons at risk for the disease.

**Table 1.** Characteristics of COVID-19 cases, stratified by vaccination status and by age group, Italy, December 2021 – May 2022.

|  |  | **12-39** | **40-59** | **60-79** | **80+** |
| --- | --- | --- | --- | --- | --- |
| **UV<sup>a</sup>** | **Diagnoses** | 3,281,968 | 2,863,586 | 979,765 | 205,400 |
|  | **Hospitalizations** | 22,304 | 37,161 | 62,869 | 46,249 |
|  | **ICU** | 720 | 4,633 | 10,585 | 1,637 |
|  | **Deaths** | 263 | 2,236 | 12,952 | 20,536 |
| **FV-120<sup>b</sup>** | **Diagnoses** | 3,550,090 | 1,394,214 | 281,414 | 44,258 |
|  | **Hospitalizations** | 13,415 | 7,678 | 8,660 | 5,303 |
|  | **ICU** | 195 | 478 | 815 | 170 |
|  | **Deaths** | 32 | 297 | 1,449 | 1,868 |
| **FV+120<sup>c</sup>** | **Diagnoses** | 5,080,841 | 3,898,449 | 1,480,538 | 231,515 |
|  | **Hospitalizations** | 14,358 | 19,891 | 54,543 | 43,383 |
|  | **ICU** | 292 | 1,092 | 5,097 | 1,094 |
|  | **Deaths** | 165 | 989 | 9,359 | 17,084 |
| **FV+B<sup>d</sup>** | **Diagnoses** | 4,625,892 | 5,905,677 | 3,641,826 | 1,339,207 |
|  | **Hospitalizations** | 12,743 | 23,026 | 64,379 | 92,712 |
|  | **ICU** | 318 | 1,273 | 4,628 | 2,429 |
|  | **Deaths** | 117 | 759 | 7,658 | 23,522 |

<sup>a</sup>**Unvaccinated cases (UV):** all notified cases with a confirmed diagnosis of infection with SARS-CoV-2 virus who did not receive any dose of vaccine or were vaccinated with the first dose or a single dose of vaccine within 14 days before diagnosis.
<sup>b</sup>**Cases of individuals fully vaccinated $\leq 120$ (FV-120):** all notified cases with a confirmed diagnosis of infection with SARS-CoV-2 virus beginning on day 14 after administration of the second dose and for the next 120 days.
<sup>c</sup>**Cases of individuals fully vaccinated $> 120$ (FV+120):** all notified cases with a confirmed diagnosis of infection with SARS-CoV-2 virus made more than 120 days after the 14th day after administration of the second dose who have not received the additional/booster dose within the previous 14 days.
<sup>d</sup>**Cases of individuals fully vaccinated with additional dose (FV+B):** all notified cases with a confirmed diagnosis of infection with SARS-CoV-2 virus at least 14 days after administration of the additional dose or booster.

**Table 2.**
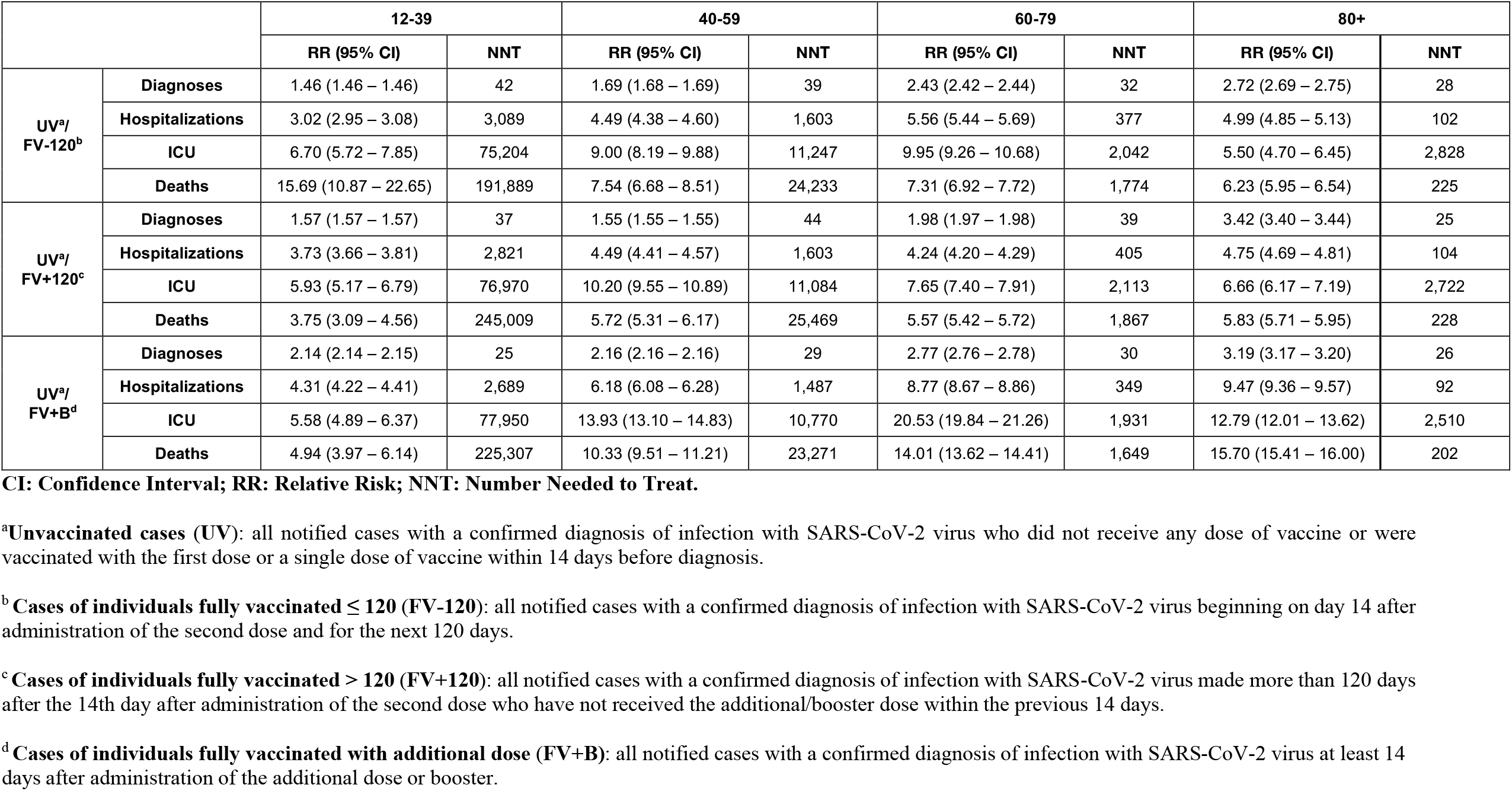
Relative risk and Number needed to treat of diagnoses and diagnoses with subsequent hospitalization and death among COVID-19 cases, stratified by vaccination status and by age group, Italy, December 2021 – May 2022.

Figure 1 describes the IR per 100,000 persons stratified by vaccination status and by age group. During the period under analysis, rates of COVID-19 cases were lowest among fully vaccinated persons with a booster dose, compared with fully vaccinated persons without a booster dose, and much lower than rates among unvaccinated persons aged ≥ 60 years.

**Figure 1.**
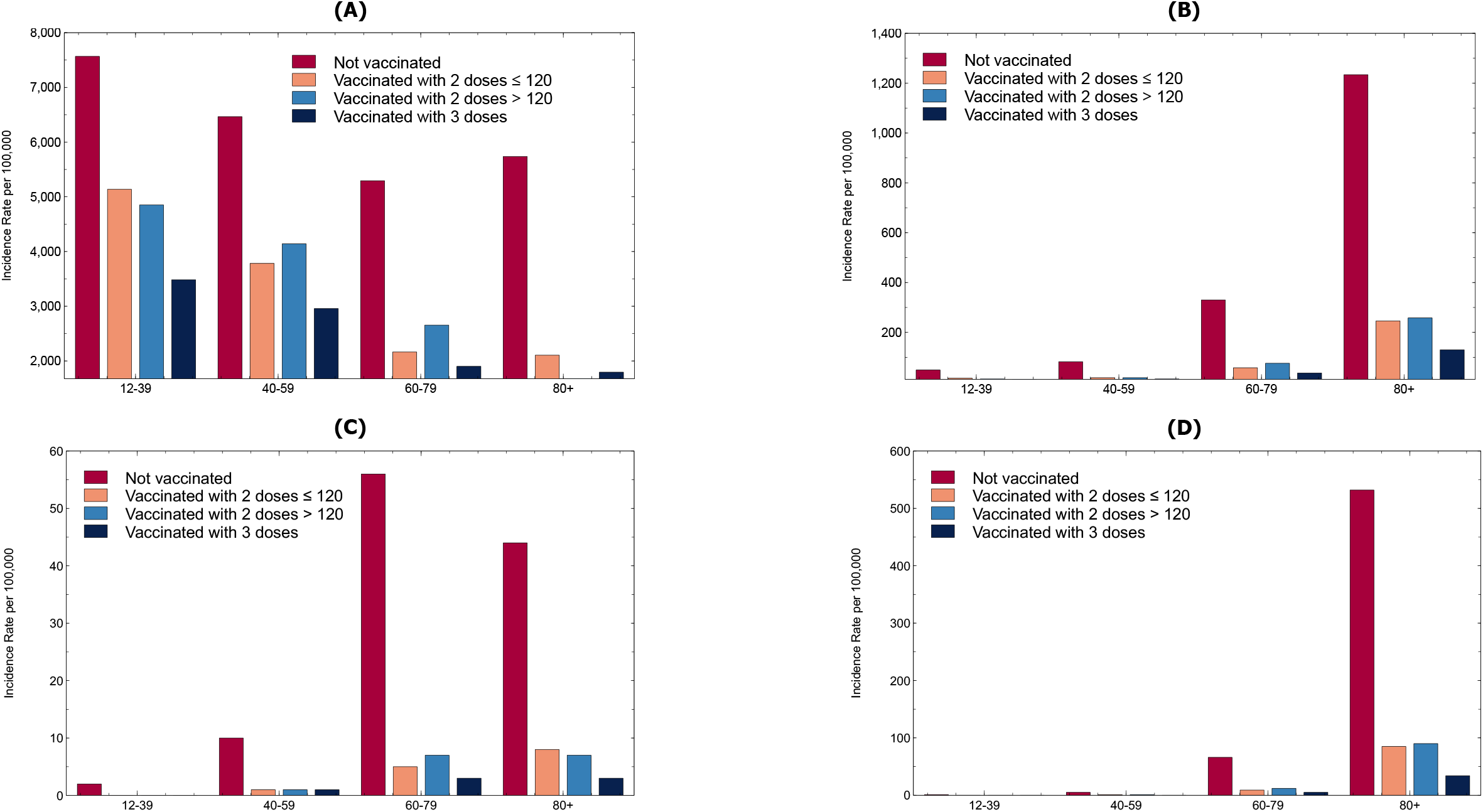
Incidence rate of COVID-19 diagnoses (A), and diagnoses with subsequent hospitalization (B), ICU admission (C), and death (D) by vaccination status and by age group.

Specifically, unvaccinated persons aged 60-79 years (330 hospitalizations per 100,000) and persons aged ≥ 80 years (1,234 hospitalizations per 100,000) had 8.77 (95% CI: 8.67-8.86) and 9.47 (95% CI: 9.36-9.57) times the risks for infection and COVID-19–associated hospitalization, respectively, compared with fully vaccinated persons who received booster doses, and 4.11 (95% CI: 4.02-4.19) and 5.16 (95% CI:5.05-5.27) times the risks compared with fully vaccinated persons without booster doses (see Supplementary Figure S2).

Among persons hospitalized with COVID-19, the risks of admissions to ICU were higher in the unvaccinated persons aged 60-79 years (IR: 56 admissions to ICU per 100,000; RR: 20.53 (95% CI: 19.84-21.26)) followed by persons aged ≥ 80 years (IR: 44 admissions to ICU per 100,000; RR: 12.79 (95% CI: 12.01-13.62)), compared with fully vaccinated persons who received booster doses (see Supplementary Figure S3).

Considering the COVID-19–associated deaths, the relative risk was highest for unvaccinated persons aged ≥ 80 years (RR=15.70 (95% CI: 15.41-16.00) times higher than for those vaccinated with booster dose, see Supplementary Figure S4), with IR reaching a value of 532 per 100,000 persons.

## Discussion

This work suggests that in Italy the vaccination with additional/booster dose considerably reduced the risk at all ages of a COVID-19 diagnosis and subsequent hospitalization and death (see Supplementary Figure S5 and Supplementary Figure S6). As of 18 May 2022, the vaccine effectiveness (VE) of vaccinated persons with 2 doses within 90 days in preventing infection was 44.0% (95% CI: 43.8%-44.1%), i.e., there was a risk reduction of about 44% for those vaccinated within 90 days compared to the unvaccinated. Between 90 and 120 days after the administration of the second dose, the estimated VE in preventing diagnoses was 33.5% (95% IC: 33.3%-33.7%), rising to 45.9% (95% IC: 45.8%-46.0%) after 120 days, and 57.6% (95% IC: 57.5%-57.7%) in individuals with additional/booster dose.

In the case of severe disease (see Figure 2(B)), VE for vaccinated persons with 2 doses within 90 days, between 91 and 120 days and over 120 days was 70.8% (95% IC: 69.9%-71.8%), 69.6% (95% IC: 68.3%-70.8%) and 71.5% (95% IC: 71.0%-72.0%) respectively, whereas it was 87.8% (95% IC: 87.6%-88.0%) in vaccinated with an additional/booster dose.

**Figure 2.**
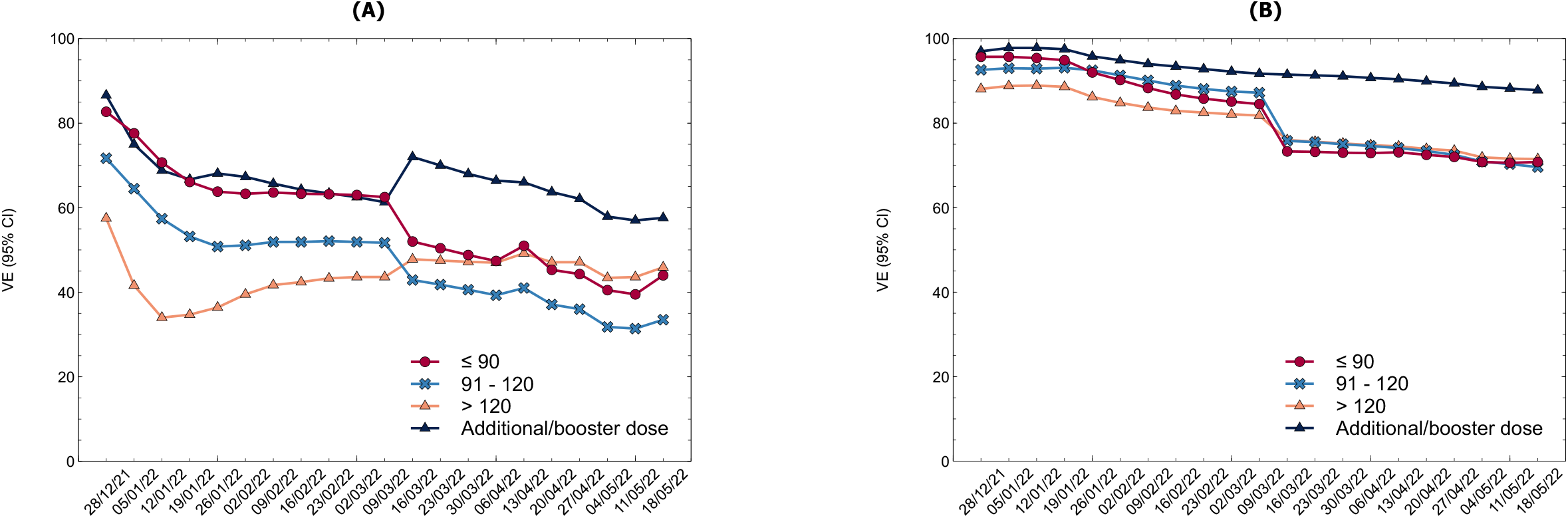
Vaccine effectiveness (VE) against SARS-CoV-2 infections (A), and severe disease (B), by vaccination status.

The results of this work have at least three limitations. First, comparisons of VE estimates between age groups must be interpreted with caution because of differences in the timing of vaccine availability and predominant variants when the vaccine became available for different age groups. Second, VE was not assessed by vaccine product (e.g., Comirnaty, Spikevax) due to lack of data. Third, it was not possible to distinguish whether a third dose was received as an additional dose to complete the primary vaccine cycle for immunocompromised persons or as a booster dose after completion of the primary vaccine cycle to assess VE in the two categories.

## Conclusion

The results underscore the importance of receiving a third dose of COVID-19 mRNA vaccine to prevent both infection and severe COVID-19, especially when the effectiveness of 2 doses is significantly reduced against the Omicron variant. It is recommended that persons aged ≥ 60 years who have received the second dose of COVID-19 vaccine should receive a third dose when eligible. Moreover, all unvaccinated persons should get vaccinated as soon as possible.

## Data Availability

All data produced in the present study are extracted from the bulletins of the Istituto Superiore di Sanit&agrave (ISS). More details are available at https://github.com/fbranda/INFN-ISS/tree/main/Report_ISS/Efficacia_vaccinale

## Footnotes

1 https://www.epicentro.iss.it/coronavirus/bollettino/Bollettino-sorveglianza-integrata-COVID-19_28-dicembre-2021.pdf

2 https://github.com/fbranda/INFN-ISS/tree/main/Report_ISS/Efficacia_vaccinale

