## Supplementary material for "Impact of the additional/booster dose of COVID-19 vaccine against severe disease during the epidemic phase characterized by the predominance of the Omicron variant in Italy, December 2021 - May 2022"

### 2x2 Table and event rates

In a study measuring dichotomous events, all effectiveness measures are derived from the 2x2 table showing the number of patients in the two groups (treatment and control) in relation to the presence/absence of the event.

| Group | Event YES | Event NO |
|---|---|---|
| Treatment | a | b |
| Control | c | d |

**Supplementary Table S1. 2x2 Table**

To evaluate the effectiveness of a vaccine, event rates are calculated, i.e., the incidence of events in the two groups under analysis:

- **Experimental Event Rate** (**EER = a/a+b**), i.e., the incidence of the event in the treatment group
- **Control Event Rate** (**CER = c/c+d**), i.e., the incidence of the event in the control group

By combining EER and CER, we obtain the following measures of effectiveness[1]:

1. **Relative measures of effectiveness**

    - **Relative Risk Reduction** (**RRR**) expresses the proportional reduction of the risk of the event in the treatment group compared to the control group; it is expressed as a percentage value.
    - **Relative Risk** (**RR**) measures the risk of the event in the treatment group compared to the control group; it is expressed in decimal values.
    - **Odds Ratio** (**OR**) is the ratio of the probability of the event in patients in the treatment group to the probability of the event in patients in the control group patients; it is expressed in decimal values. The OR value can be considered superimposable on the RR when the CER is low (< 10%); however, as the CER increases, the OR moves progressively away from the RR, leading, for CER values above 15-20%, to a further overestimation of treatment efficacy. For this reason, it is not often used.

[1] Cartabellotta A. RRR, RR, OR, ARR, NNT: che confusione!. Istruzioni per l’uso. 2012. Available at: https://www.jamd.it/wp-content/uploads/2017/02/2012_3_10.pdf

2. **Absolute measures of effectiveness**

- **Absolute Risk Reduction** (**ARR**) expresses the absolute reduction in the risk of the event in the treatment group compared to the control group. Since it is derived from the CER-EER formula, the ARR is also called *risk difference* and is generally expressed in decimal values. ARR is a difficult measure to interpret and to transfer to clinical decisions, because the quantitative value is so small that it underestimates the effectiveness of the treatment.

- **Number Needed to Treat** (**NNT**) indicates the number of patients to be treated to prevent an event. It is ideal for reporting trial results because it expresses a patient-related whole number that is easy to interpret and transfer to clinical decision-making. The ideal NNT is 1, where everyone improves with treatment, and no one improves with control. The higher the NNT value, the less effective the treatment.

| |
|---|
| RRR = [CER - EER]/CER |
| RR = EER/CER |
| OR = [EER/1 - EER] / [CER/1 - CER] |
| ARR = CER-EER |
| NNT = 1/ARR |

**Supplementary Table S2. Summary of the main measures for evaluating vaccine effectiveness.**

**Example:** Assuming the vaccine effectiveness was tested on a population of 34,922 people, 17,411 vaccinated and 17,511 received the placebo. The total number of Covid-19 cases (during the 45-day observation period) was 170, of which 8 among the vaccinated and 162 among the unvaccinated. The risk of contracting the disease during the time considered takes on the following values in the two groups compared:

| | **COVID-19** | **Healthy** | **Total** |
|---|---|---|---|
| **Vaccinated** | 8 | 17,403 | 17,411 |
| **Placebo** | 162 | 17,349 | 17,511 |
| **Total** | 170 | 34,752 | 34,922 |

- Risk for the vaccinated: $R_V$: 8/17.411=0,046%

- Risk for the unvaccinated: $R_{NV}$: 162/17.511=0,925%

The relative risk or risk ratio (RR) indicates that the unvaccinated have a risk of contracting the disease that is about 20 times greater than those who have received the vaccine:

$$RR_{NV/V} = 0,925\% / 0,046\% = 20,134$$

A vaccinated individual, on the other hand, has a 5% higher risk of becoming infected and ill than an unvaccinated person:

$$RR_{V/NV} = 0,046\% / 0,925\% = 0,04967$$

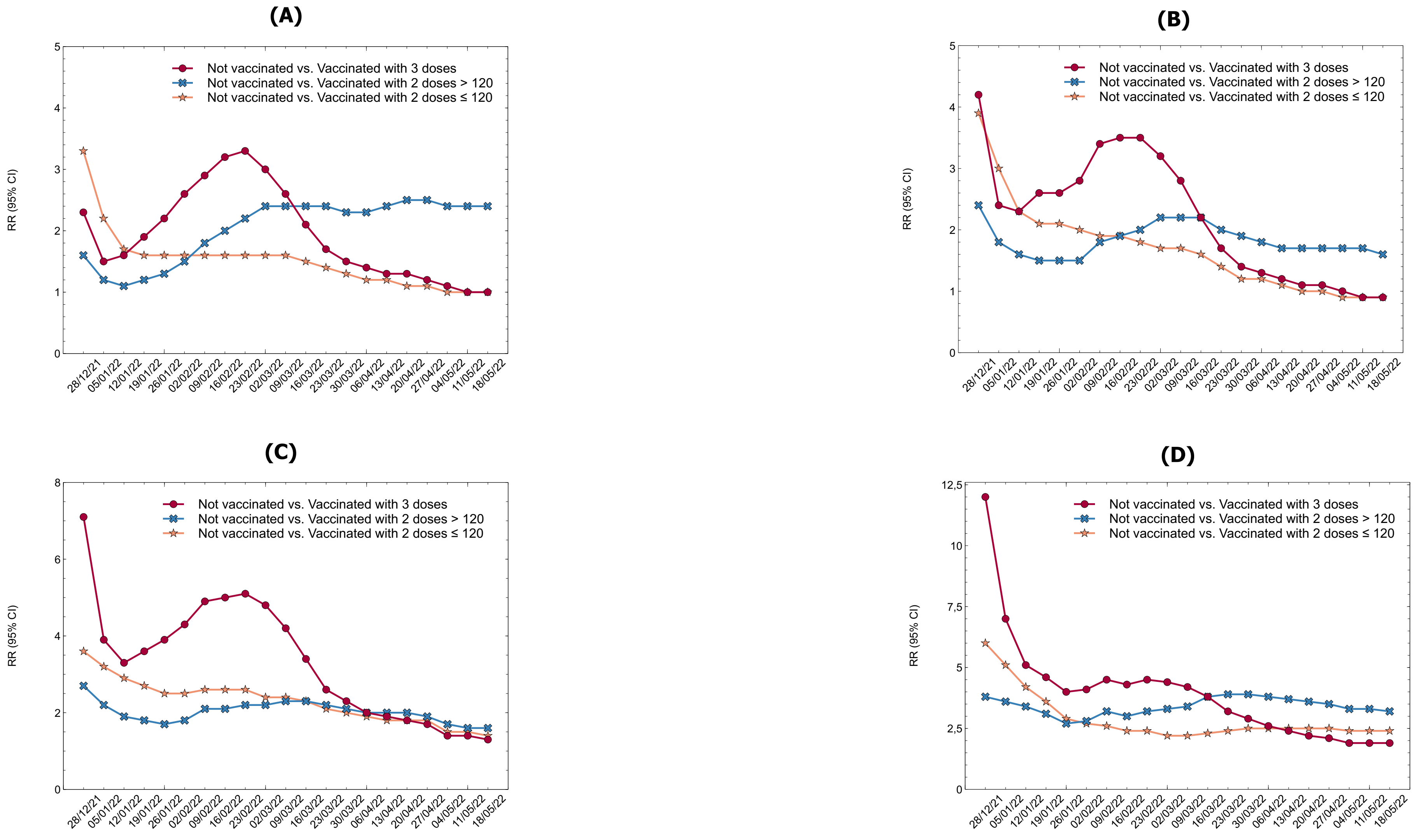


**Supplementary Figure S1. Relative Risk (RR) of any COVID-19 diagnosis (symptomatic or asymptomatic) by vaccination status and by age group. (A) Persons 12-39 years of age; (B) Persons 40-59 years of age; (C) Persons 60-79 years of age; (D) Persons 80+ years of age.**

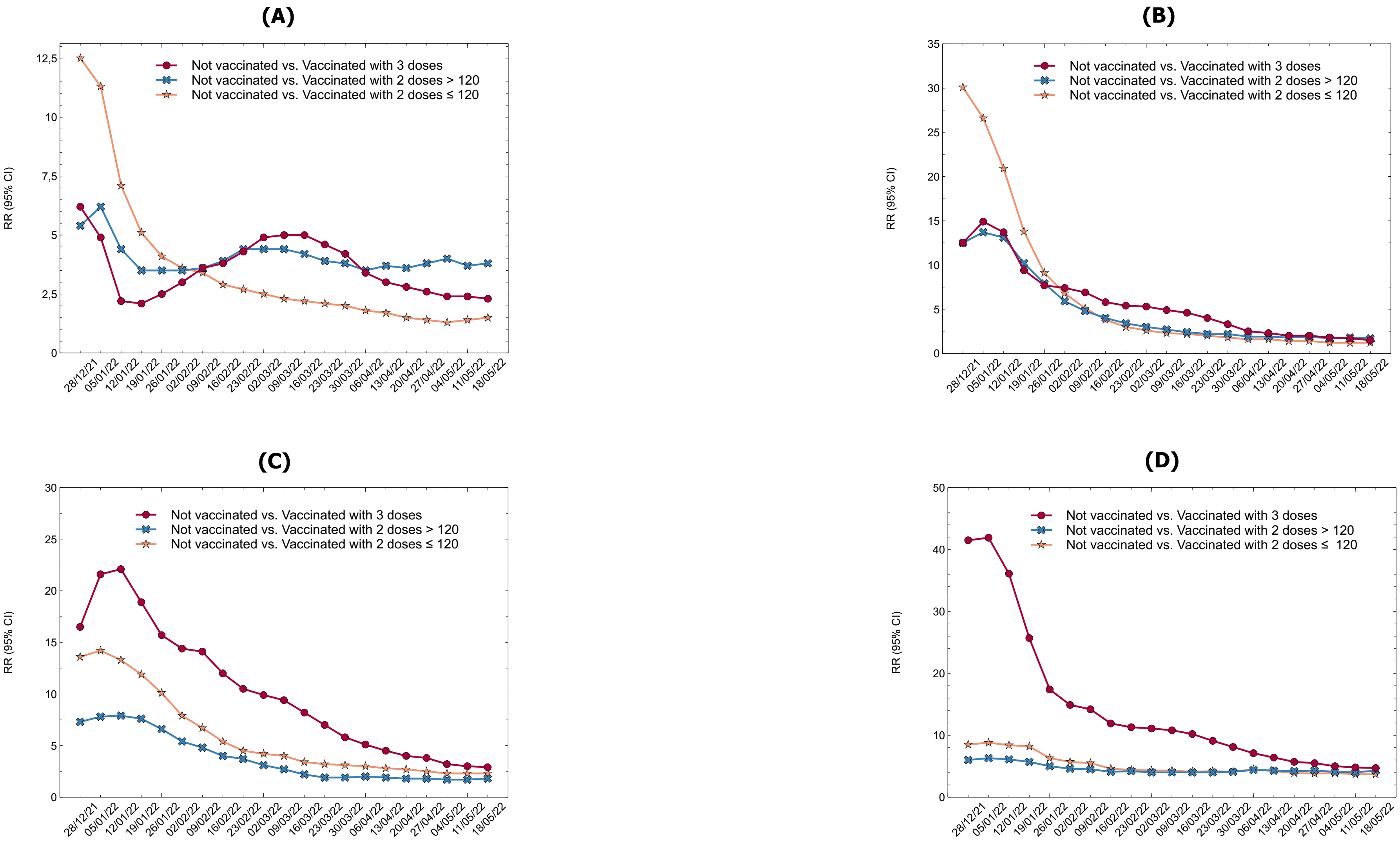


**Supplementary Figure S2. Relative Risk (RR) of diagnoses with subsequent hospitalization, by vaccination status and by age group. (A) Persons 12-39 years of age; (B) Persons 40-59 years of age; (C) Persons 60-79 years of age; (D) Persons 80+ years of age.**

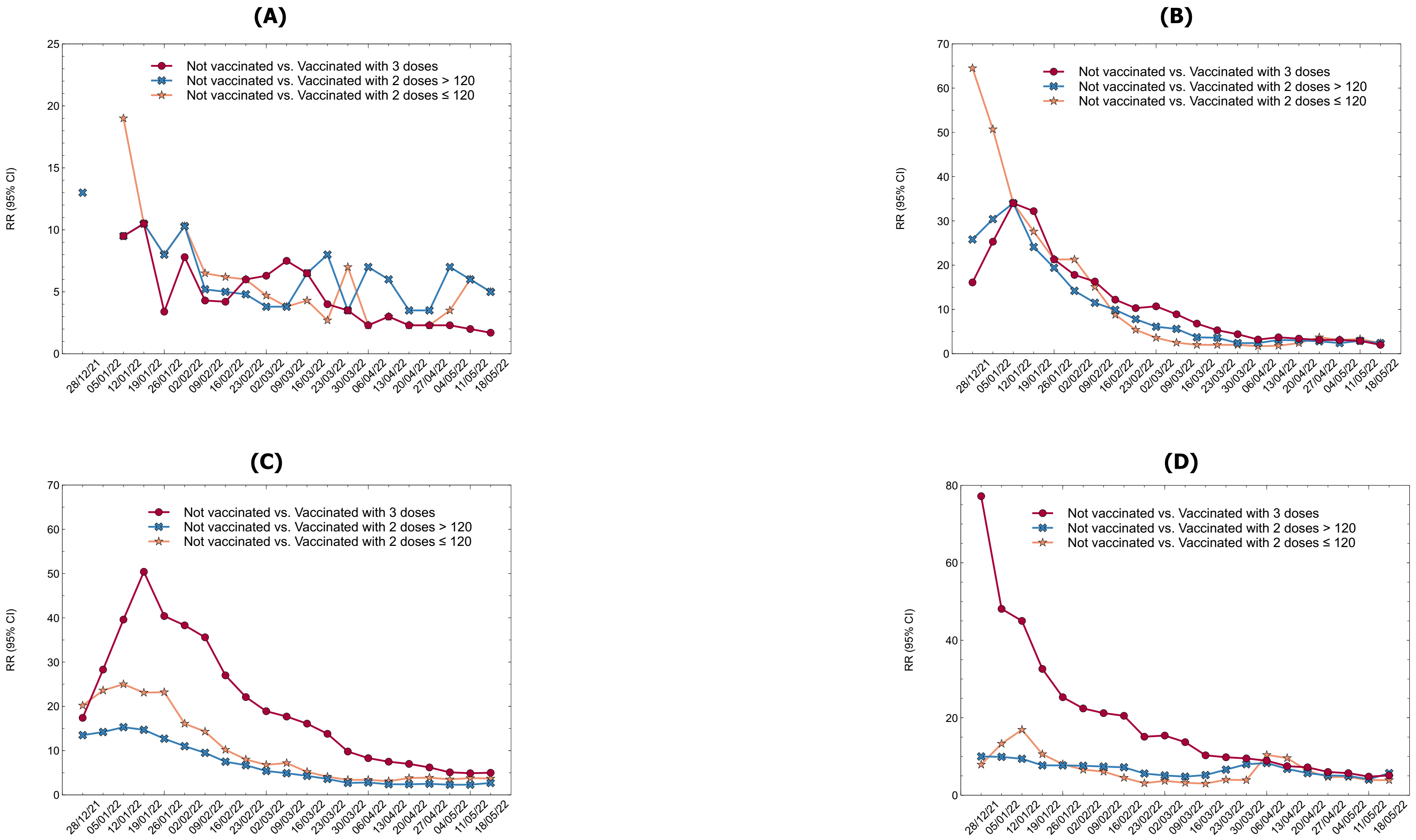


**Supplementary Figure S3. Relative Risk (RR) of diagnoses with subsequent admission to ICU, by vaccination status and by age group. (A) Persons 12-39 years of age; (B) Persons 40-59 years of age; (C) Persons 60-79 years of age; (D) Persons 80+ years of age.**

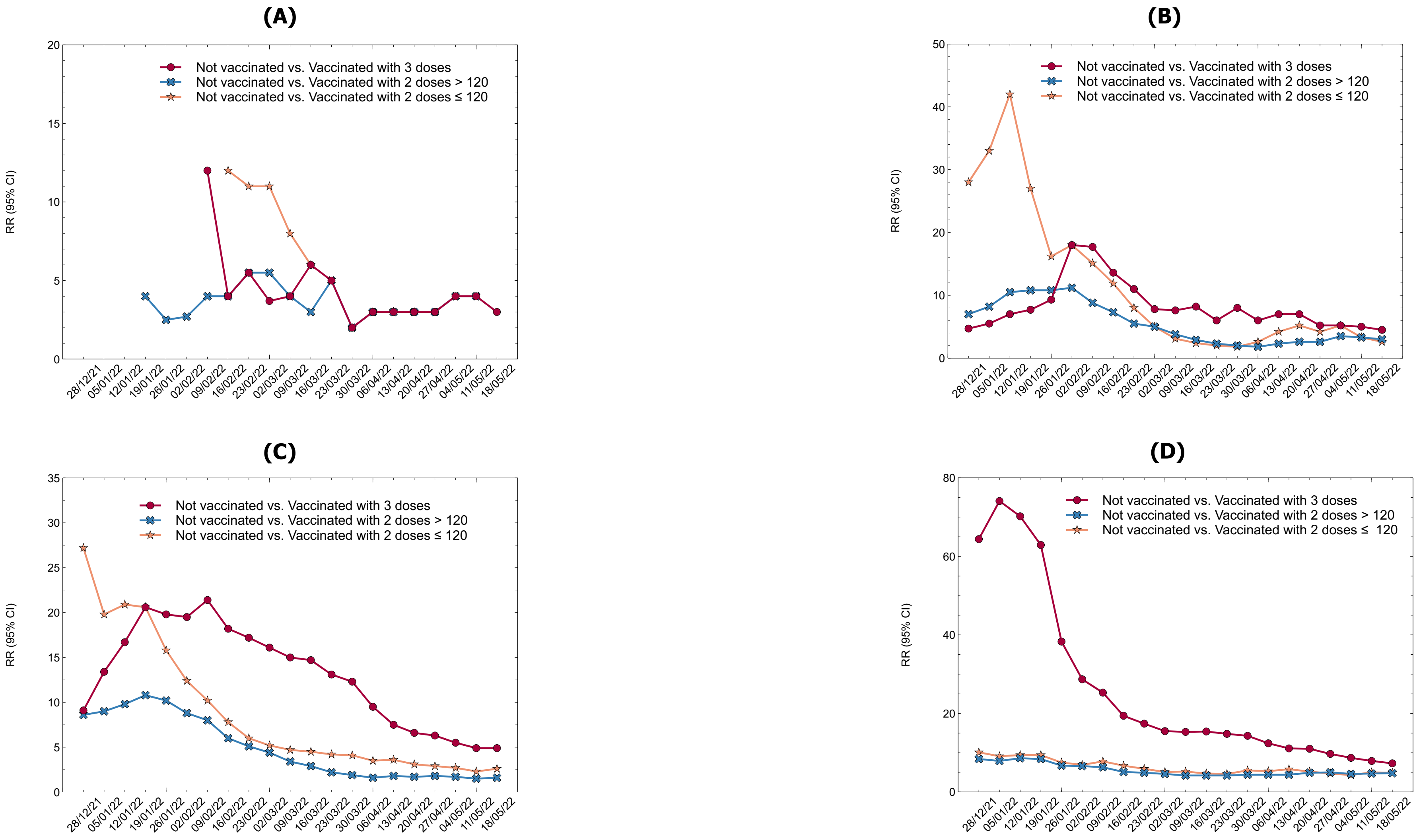


**Supplementary Figure S4. Relative Risk (RR) of diagnoses with subsequent death, by vaccination status and by age group. (A) Persons 12-39 years of age; (B) Persons 40-59 years of age; (C) Persons 60-79 years of age; (D) Persons 80+ years of age.**

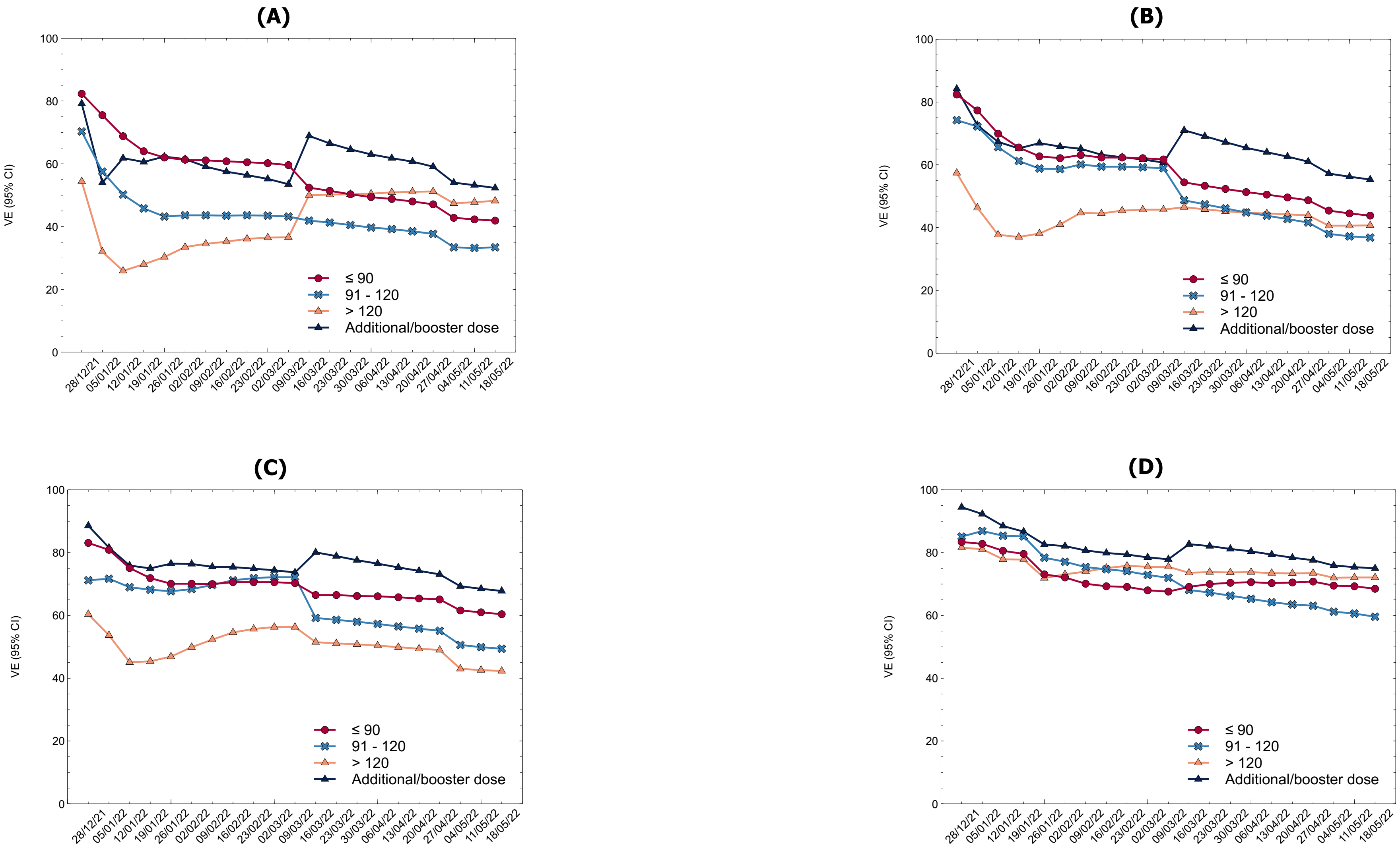


**Supplementary Figure S5. Vaccine effectiveness (VE) against SARS-CoV-2 infections, by vaccination status and by age group. (A) Persons 12-39 years of age; (B) Persons 40-59 years of age; (C) Persons 60-79 years of age; (D) Persons 80+ years of age.**

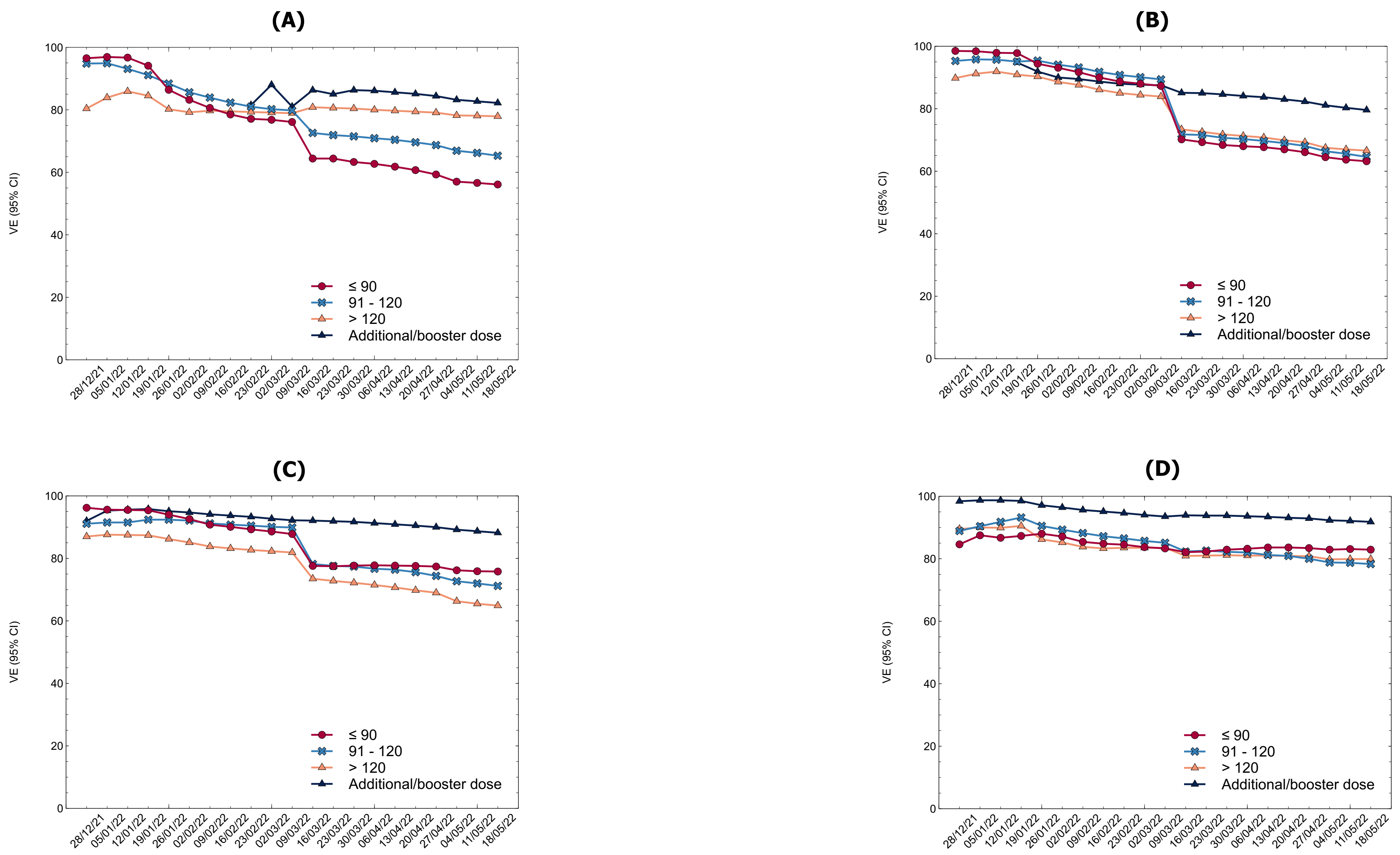


**Supplementary Figure S6. Vaccine effectiveness (VE) against severe COVID-19, by vaccination status and by age group. (A) Persons 12-39 years of age; (B) Persons 40-59 years of age; (C) Persons 60-79 years of age; (D) Persons 80+ years of age.**
